# A Susceptible-Infected-Removed (SIR) model of COVID-19 epidemic trend in Malaysia under Movement Control Order (MCO) using a data fitting approach

**DOI:** 10.1101/2020.05.01.20084384

**Authors:** Wan Nor Arifin, Weng Howe Chan, Safiya Amaran, Kamarul Imran Musa

**Author notes:** These authors contributed equally to this work (Wan Nor Arifin, Weng Howe Chan, Safiya Amaran).

## Abstract

**Background:** In this work, we presented a Susceptible-Infected-Removed (SIR) epidemiological model of COVID-19 epidemic in Malaysia post- and pre-Movement Control Order (MCO). The proposed SIR model was fitted to confirmed COVID-19 cases from the official press statements to closely reflect the observed epidemic trend in Malaysia. The proposed model is aimed to provide an accurate predictive information for decision makers in assessing the public health and social measures related to COVID-19 epidemic.

**Methods:** The SIR model was fitted to the data by minimizing a weighted loss function; the sum of the residual sum of squares (RSS) of infected, removed and total cases. Optimized beta (β),), gamma (γ) parameter values) parameter values and the starting value of susceptible individuals (*N*) were obtained.

**Results:** The SIR model post-MCO indicates the peak of infection on 10 April 2020, less than 100 active cases by 8 July 2020, less than 10 active cases by 29 August 2020, and close to zero daily new case by 22 July 2020, with a total of 6562 infected cases. In the absence of MCO, the model predicts the peak of infection on 1 May 2020, less than 100 active cases by 14 February 2021, less than 10 active cases by 26 April 2021 and close to zero daily new case by 6 October 2020, with a total of 1.6 million infected cases. **Conclusion**: The results suggest that the present MCO has significantly reduced the number of susceptible population and the total number of infected cases. The method to fit the SIR model used in this study was found to be accurate in reflecting the observed data. The method can be used to predict the epidemic trend of COVID-19 in other countries.

## Introduction

The novel coronavirus disease (COVID-19) caused by SARS-CoV-2 was announced as a pandemic on 11 March 2020 by the World Health Organization, less than 3 months since the epidemic started in Wuhan, China. As of 29 April 2020, the number of cases climbed above 3 million with a death toll of over 200,000 worldwide [1, 2]. In response to COVID-19, many countries implemented a large scale public health and social measures (PHSM), including movement restrictions, closure of schools and businesses, geographical area quarantine, and international travel restrictions. These measures are sometimes referred to as “lockdown” or “*cordon sanitaire*”.

Similarly, in Malaysia a large scale PHSM called Movement Control Order (MCO) was announced to the whole country under the Prevention and Control of Infectious Diseases Act 1988 (Act 342) on 16 Mac 2020. Following the MCO, all gatherings, whether for religious, sports, recreational, social or cultural purposes are banned. All universities, schools and non-essential sectors are closed. Journeys from one place to another within any infected local area are not allowed except for the following purposes: to perform any official duty; to make a journey to and from any premises providing essential services, non-essential services or food supply; to purchase, supply or deliver food or daily necessities; to seek healthcare or medical services; or any other special purposes as may be permitted by the Director General of Health. Inter-state journeys from one infected local area to another infected local area are not allowed unless prior written permission of a police officer in charge of a police station is obtained [3].

The first MCO (MCO1) started on 18 March until 31 March 2020, for a duration of two weeks. It was then continued every 2 weeks, second MCO (MCO2) till 14 April 2020, third MCO (MCO3) till 28 April 2020 and fourth MCO (MCO4) till 12 May 2020. Malaysia is still under MCO until the day of writing [2].

PHSM involving the whole country continuously for almost 8 weeks is not without a burden. It has a major social consequences and economic costs. At the moment, the decision to implement MCO was acceptable to many in view of disease casualties, but the question is how long should the MCO be in place? There is a need for Malaysian authority to assess and balance the benefits and potential harms of adjusting these MCO duration, so as not to trigger a resurgence of COVID-19 cases and jeopardize the health of the population.

In this study, we presented a Susceptible-Infected-Removed (SIR) epidemiological model of the COVID-19 epidemic in Malaysia following the MCO and prior to the MCO. To make sure the model closely reflects the observed epidemic trend, the proposed SIR model was fitted to confirmed COVID-19 cases from the official press statements by the Director General of Health, Malaysia by optimizing a chosen loss function. The proposed model is aimed to provide an accurate predictive information in assessing the public health and social measures for COVID-19 epidemic and in due course make timely future plans.

## Methods

### Data Sources

This study utilized the COVID-19 data set from https://wnarifin.github.io/covid-19-malaysia/, which is updated daily by WNA until the date of writing. The data set contains the number of new and cumulative cases, deaths and recovered cases, together with the number of active cases in ICU and respiratory support. The data set was initially setup based on Our World in Data COVID-19 data set (https://covid.ourworldindata.org/data/ecdc/full_data.csv). Because of a number of discrepancies between the official reports and the data set from Our World in Data, a corrected and updated data set was created by WNA according to the official daily press statements by the Director General of Health Malaysia (https://kpkesihatan.com/).

### SIR Model

One of the mathematical models that describe the population level dynamics of infectious diseases is the classical SIR compartmental disease model, which is based on the work of Kermack and Kendrick in 1930s [4]. Conceptually, the SIR model is presented schematically as shown in Figure 1 below:

**Figure 1:**
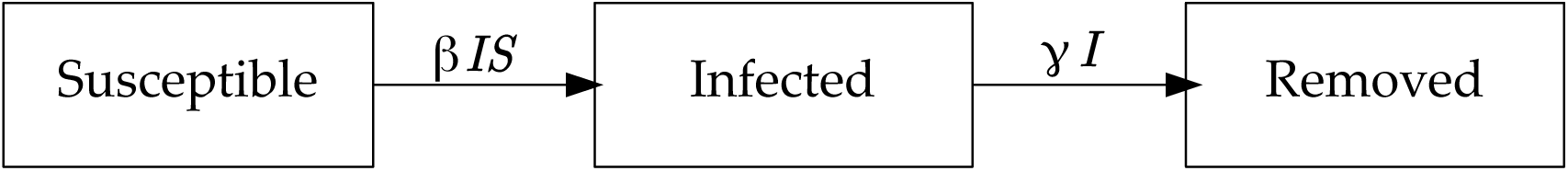
Susceptible, Infected and Removed compartments in the SIR model.

Assuming zero birth and death rate, this SIR model can be defined by a set of ordinary differential equations as follows [4, 5]:

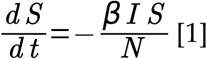

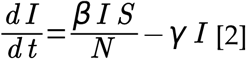

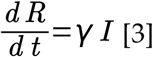

where *S =* susceptible individuals, *I =* infectious individuals (active cases), *R =* removed individuals (recover and death), and *N = S(t) + I(t) + R(t)*. 1/*γ* is the time until recovery and β), is the transmission rate (contact rate x probability of transmission given contact).

The basic reproductive number, *R*_0_ is defined as the “average number of secondary cases arising from a typical primary case in an entirely susceptible population” [4]. The *R*_0_ is given as:

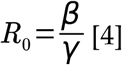

### Finding Optimal Model Parameters

To fit the model to the data, the following weighted loss function was defined, taking the sum of the residual sum of squares (RSS) for *ϒ* = *I*, *R* and total cases (*I + R*):

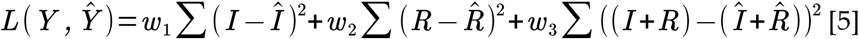

The model was fitted to the data using the methods described in [6, 7] in R software environment [8], utilizing deSolve R package [9]. For optimization procedure, the parameters value were set within clinically reasonable start, upper and lower limits: β = {1/ 2, 1/ 100, 1} and γ = {1/14, 1/42 (6 weeks [10]), 1/11 (2 weeks [10], minus 3 days to allow parameter exploration)}.

Apart from finding the β and γ parameter values, in Malaysian context, experts were divided about the initial number of susceptible individuals, *S*, which is *N*. Thus, we also estimated the *N* of the model by iterating over a decreasing number of *N*, starting from *N =* 32.68 million of Malaysian population in 4^th^ quarter of 2019 [11]. The optimal *N* was the number that fitted within the SIR model with the smallest value of the loss function. The pseudo-code for the iteration procedure is given in Algorithm 1 below. In our experiment, Step = MaxStep = 7 was found to be sufficient.

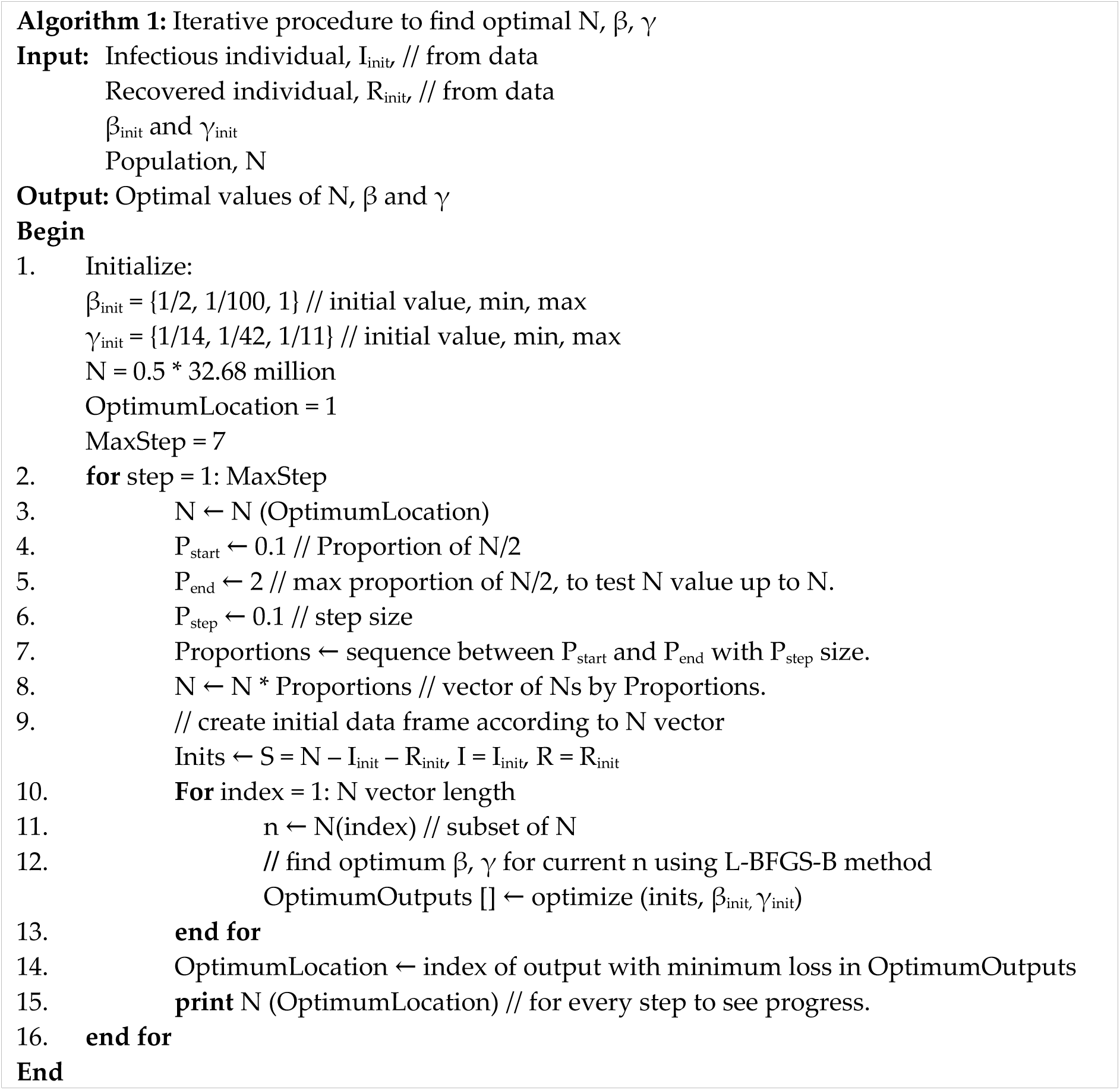

Once the optimal β, γ and *N* values that minimized the loss function were found, projected data were obtained from the SIR model using the optimized parameters. Using the data, the SIR model was plotted using ggplot2 R package [12]. The goodness of fit and error rate measures of the fitted model were given in form of R^2^ and mean absolute percentage error (MAPE).

## Results

For the analysis, we divided the analysis into two distinct periods: post-MCO (25 March 2020 until 29 April 2020) and pre-MCO (1 March 2020 to 24 March 2020). The post-MCO period starts one-week following the MCO initiation (18 March 2020) to allow a one-week grace period for the population to reach an optimal compliance of the MCO. We excluded imported cases from the data points by 24 April 2020 onwards as the daily new cases included a notable number of imported cases, which violated our assumption for the SIR model of no addition to the starting population. The pre-MCO period includes the first week of MCO1 as we assumed that the epidemiological pattern prior to MCO implementation is similar to the first week of MCO1. The post-MCO period analysis reflects the SIR modeling of COVID-19 epidemic trend in the presence of MCO, while the pre-MCO analysis reflects the SIR modeling of the epidemic trend in the absence of MCO.

Figures 1 and 2 show the plots of the SIR model for the post-MCO period. Figure 2 includes additional curves, assuming 5% (Beijing) and 50% (Iceland) asymptomatic cases [13].

**Figure 1:**
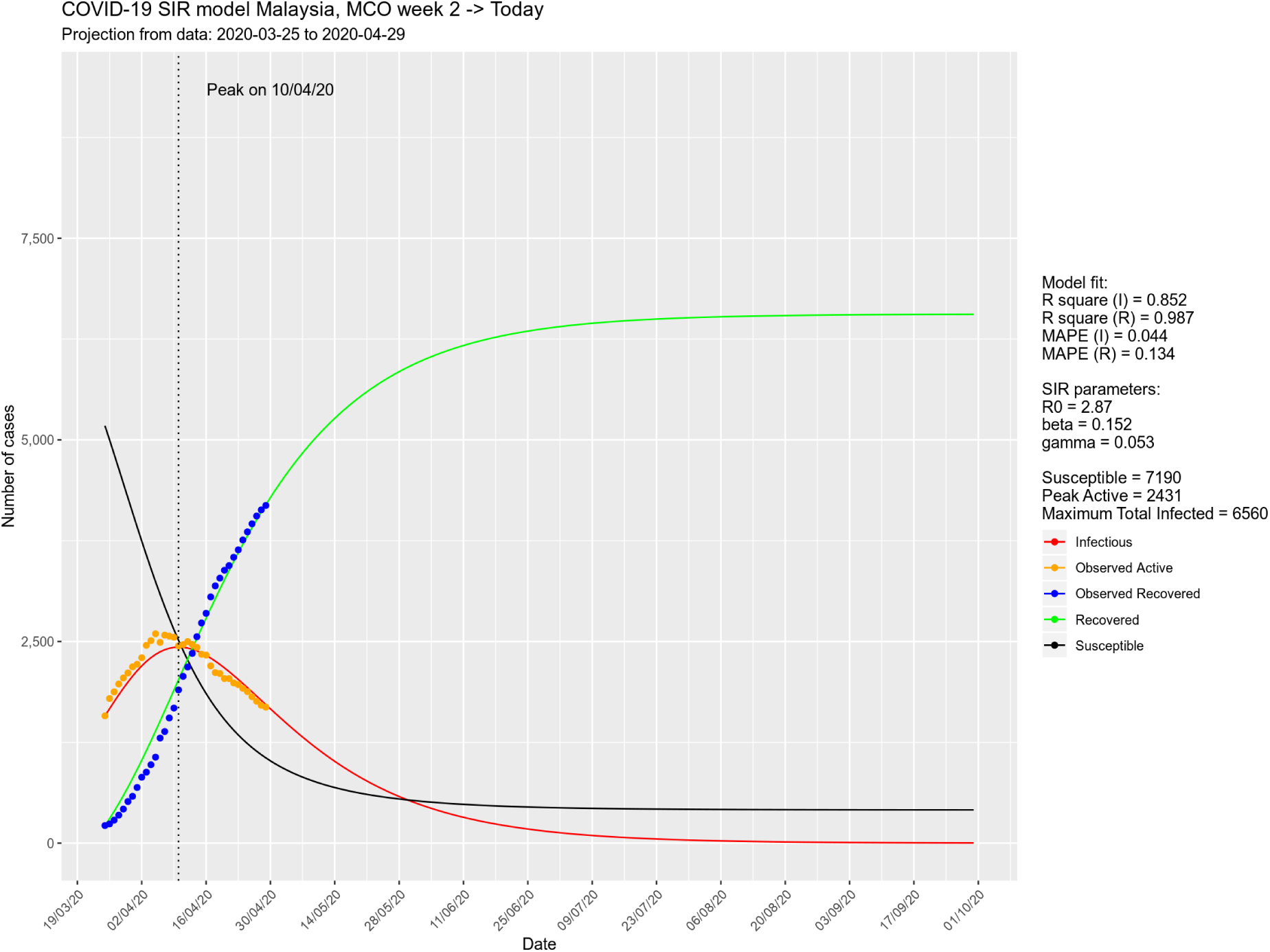
SIR model prediction for post-MCO period, based on data up to 29/4/2020.

**Figure 2:**
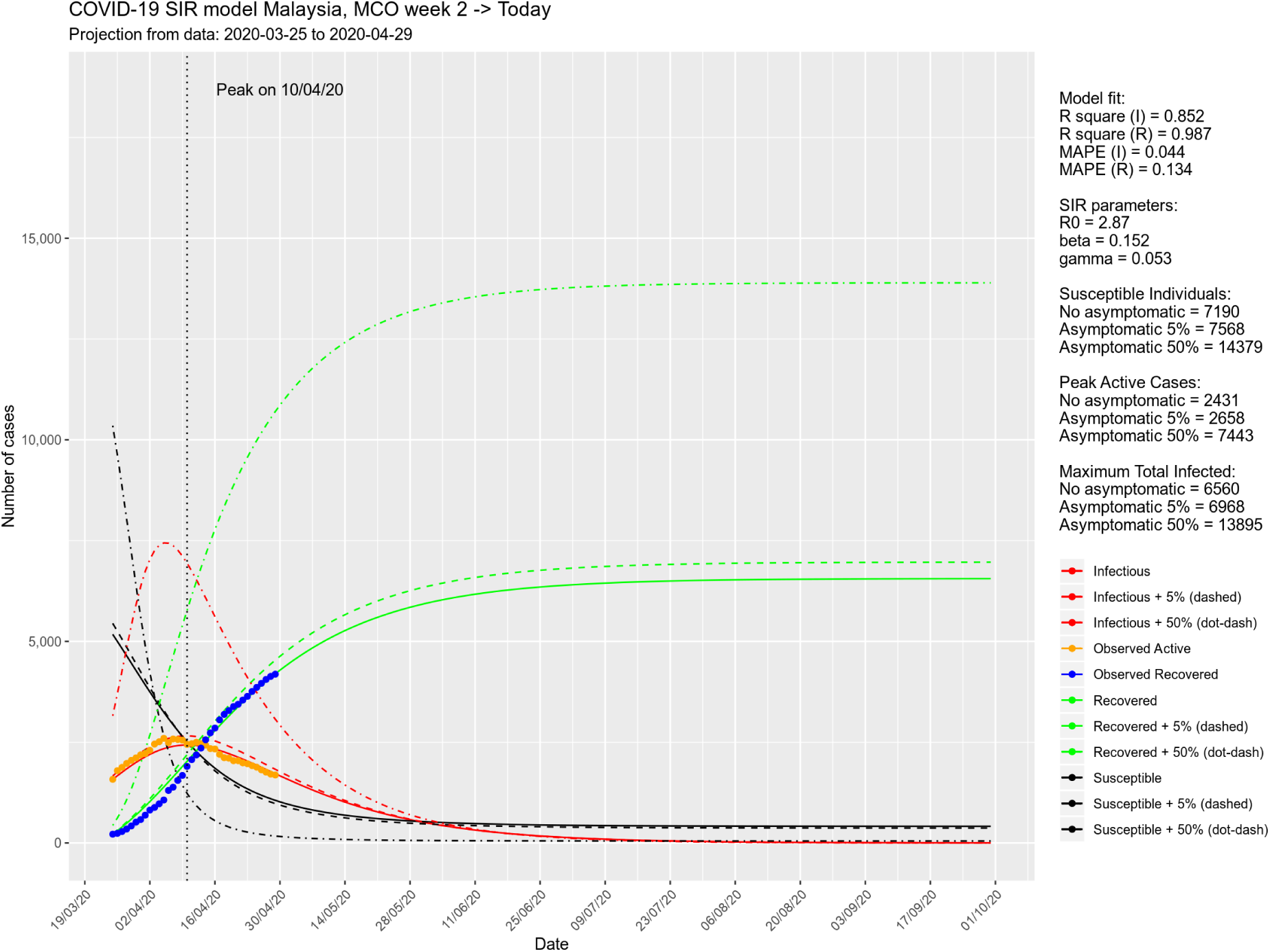
SIR model prediction for post-MCO period, based on data up to 29/4/2020, assuming 5% and 50% asymptomatic cases.

Figures 3 and 4 show the plots of the SIR model for the pre-MCO period. Figure 3 includes additional curves, assuming 5% (Beijing) and 50% (Iceland) asymptomatic cases [13]. For the pre-MCO period, the MaxStep was set to 1, which mean the loop only run once. Here, we limited the algorithm to search for the optimal parameters between 5% to 100% of the initial susceptible population so as to reflect the absence of MCO. In addition, this was also because whenever the data do not contain the peak of active cases, the algorithm is unable to find the optimal *N*.

**Figure 3:**
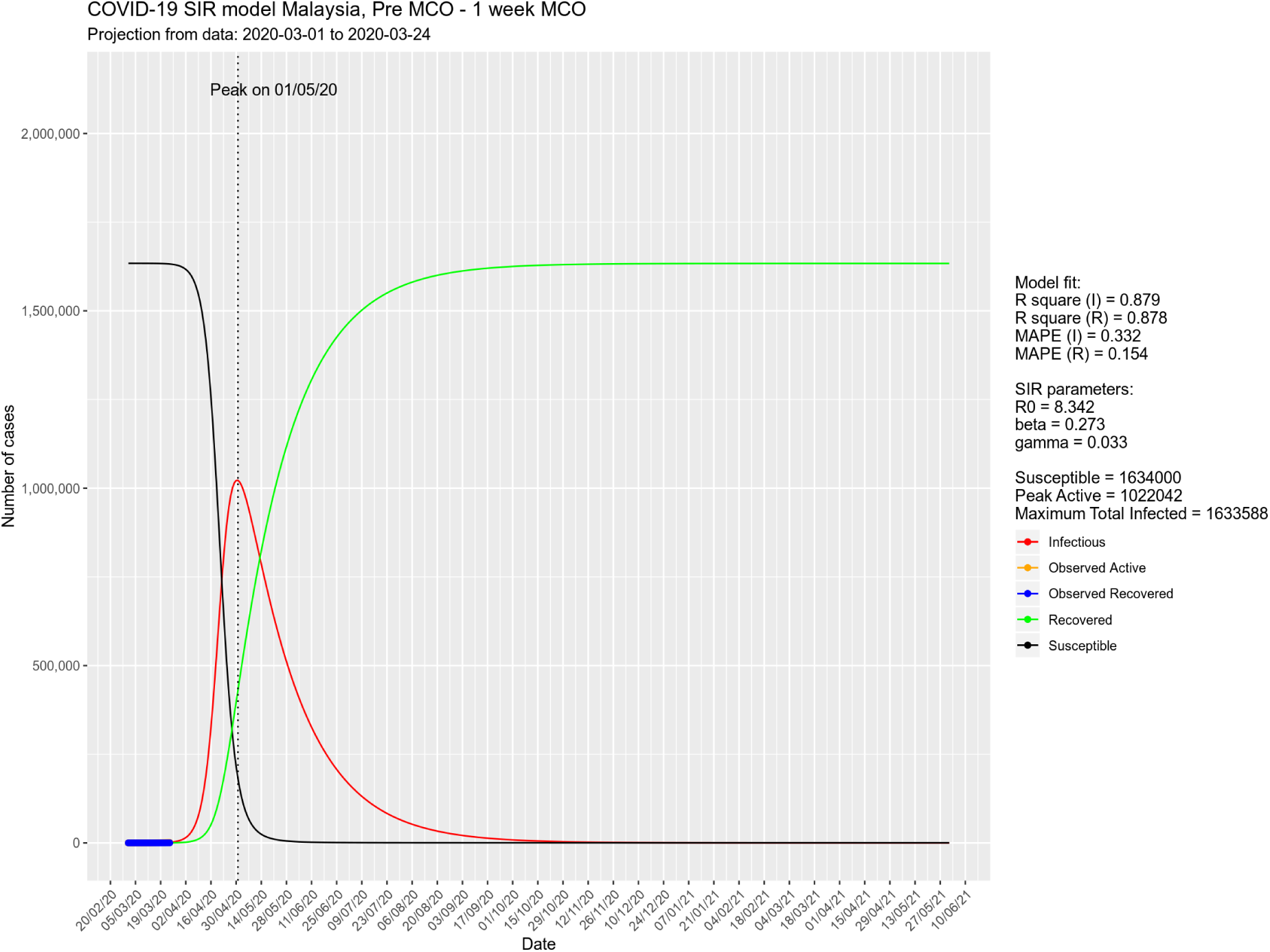
SIR model prediction based for pre-MCO period.

**Figure 4:**
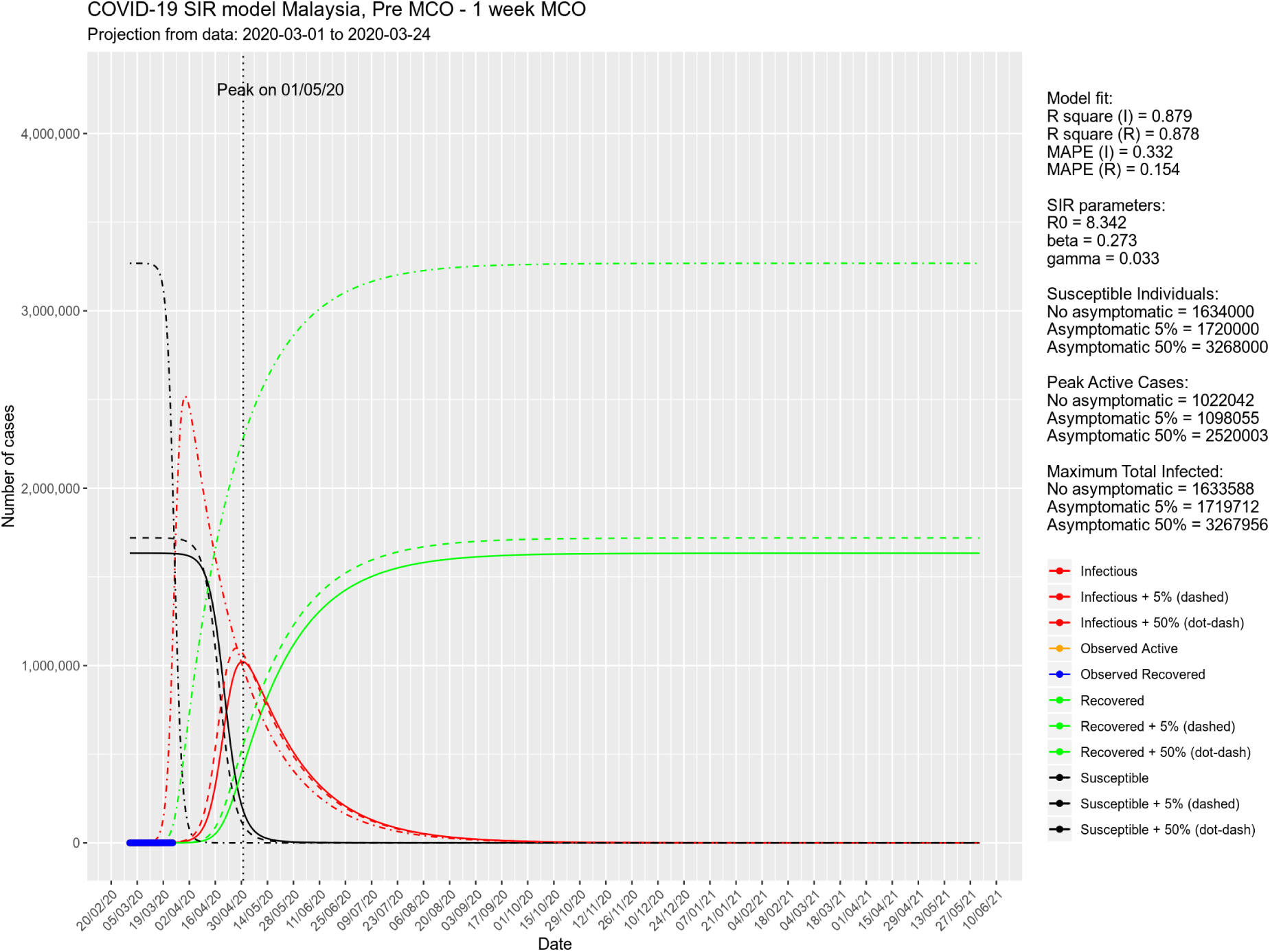
SIR model prediction based for pre-MCO period, assuming 5% and 50% asymptomatic cases.

## Discussion

The SIR model following the MCO indicates that the infection already peaked on 10 April 2020, after which the number of active cases started to decline. There will be less than 100 active cases by 8 July 2020 and less than 10 active cases by 29 August 2020, close to zero daily new case (when the predicted new case < 0.5) by 22 July 2020. The total number of infected cases was predicted to be between 6560 to 13895 cases. In contrast, if the MCO was not implemented, the infection will peak on 1 May 2020, and there will be less than 100 active cases by 14 February 2021, less than 10 active cases by 26 April 2021 and close to zero daily new case by 6 October 2020, with a larger number of total infected cases between 1.6 million to 3.3 million cases.

The fitted models for both periods maintained high goodness of fit, indicated by R^2^ values of 85.2% to 98.7% for *I* and *R* respectively. The error rates by MAPE were low for post-MCO period with MAPE (I) and MAPE (R) of 4.4% and 13.4% respectively. The error rates were slightly higher for pre-MCO period, with MAPE (I) and MAPE (R) of 33.2% and 15.4% respectively. This is because the data pattern during that period was relatively less well explained by the SIR model especially for *I*, and the number of data points was smaller as compared to post-MCO period (24 vs 36). This could also be due to these reasons: a) the Ministry of Health increased the test capacity, b) delay in receiving the test results due to overwhelmed laboratory capacity and c) the number of tests fluctuates on daily basis [14, 15]. However, this high error rate was balanced by high R^2^ for both *I* and *R* (87.9% and 87.8% respectively).

The R_0_ for all presented figures were relatively high as compared to other studies [16, 17, 18]. For post-MCO period, the R_0_ is close to 3, which is against the common assumption that the R_0_ should be low once health intervention measures kicks in. This is also the case with the pre-MCO R_0_, which is more than 8. However, these R_0_s for both periods were the ones that best explained the observed data. We propose that this observation might suggest the real infectiveness in the susceptible population, which is the fuel of the epidemic. Although the infectiveness of COVID-19 remains high following the MCO, the size of susceptible population reduced significantly as a result of MCO implementation. At the same time, it was found that MCO significantly reduced transmission rate by 44.3%, from β = 0.273 to β = 0.152.

### Limitations

The model was unable to consider imported cases because the size of susceptible population was assumed fixed at the start of each prediction period. Hence, another approach must be considered whenever imported cases are present. The performance measures provided in form of R^2^ and MAPE can be viewed as the goodness of fit and error measures for the training set, which may not be generalizable to future observations and there could be an issue with model overfitting [19]. However, given the limited number of available data points, all available data points were used for the model optimization. Given the urgency of the present situation, it is not reasonable to wait for more data points to be used as the test set to estimate the test goodness of fit and error measures. In our opinion, cross-validation method is also not reasonable in this situation given the small number of data points. Finally, our model used the available incidence data instead of the data based on date of onset [20]. Despite this limitation, we feel that our SIR model, to some extent, reflects the true COVID-19 epidemic trend.

## Conclusion

The results showed that the implementation of the MCO from 18 March 2020 until today has significantly reduced the COVID-19 transmission in Malaysia. The MCO effectively restricted the number of susceptible population, resulting in a smaller number of infected cases. Based on the model, the flattened epidemic curve is expected to be in July this year. The method used in this study to fit the SIR model was shown to be accurate in reflecting the observed data. This method can be used to predict the epidemic trend of COVID-19 in other countries.

## Authors’ Contributions

Conception and design: WNA, WHC; Analysis of the data: WNA, WHC; Interpretation of the data: WNA, WHC, SA, KIM; Drafting of the article: WNA, SA; Critical revision of the article for important intellectual content: WNA, WHC, SA, KIM; Final approval of the article: WNA, WHC, SA, KIM; Computational expertise: WNA, WHC; Epidemiological expertise: SA, KIM.

## Data Availability

The data set and codes used in this study are available at the provided links.

https://wnarifin.github.io/covid-19-malaysia/

https://github.com/cwenghowe/sircovid19mys

## Acknowledgments

We would like to thank the contributions and support from our research groups: COVID-19 Prediction Team (Universiti Teknologi Malaysia: Naomie Salim, Shuhaimi Mansor, Nor Erne Nazira Bazin, Ahmad Athif Mohd Faudzi, Anazida Zainal, Sharin Hazlin Huspi, Eric Khoo Jiun Hooi, Shaekh Mohammad Shithil; Universiti Sultan Zainal Abidin: Abdul Rahim Wong, Dr. Syed Hatim Noor @ Nyi Nyi Naing) and COVID-19 Epidemiology Modeling Team (Health Campus, Universiti Sains Malaysia: Mohd Azmi Suliman, Mohamad Zarudin Mat Said, Wira Alfatah Ab Ayah @ Ab Aziz, Che Muhammad Nur Hidayat Che Nawi, Afiqah Syamimi Masrani, Tengku Muhammad Hanis Tengku Mokhtar, Mohd Fadzali Bakar, Mohd Faizal Abdul Manaf, Ahmad Syakiren Mazalan; Jabatan Kesihatan Negeri Pahang: Sahrol Azmi Termizi).

## Availability of data and materials

The data set can be downloaded from https://wnarifin.github.io/covid-19-malaysia/. The codes for this work are available from https://github.com/cwenghowe/sircovid19mys.

